# DISCERN: A Clinical Impact-aware Framework for Radiology Report Comparison

**DOI:** 10.64898/2026.05.26.26353612

**Authors:** Rakesh Sharma, Cameron Beeche, Jessie Dong, Richard Zhuang, Huaizhi Qu, Ruichen Zhang, Vineeth Gangaram, Pulak Goswami, Jiayi Xin, Jenna Ballard, Jeffery Duda, Charles E. Kahn, Ari Goldberg, Hersh Sagreiya, Qi Long, Tianlong Chen, Walter Witschey

## Abstract

The surge in medical imaging has spurred the development of vision-language models (VLMs) to alleviate radiologist workloads. However, clinical deployment is hindered by the lack of meaningful evaluation frameworks. Current metrics - ranging from semantic similarity to large language model (LLM) based judges - often fail to distinguish between clinically trivial and critical discrepancies, poorly reflecting real-world clinical judgment. To address this, we introduce DISCERN (Discordance and Significance-aware Entity-level Radiology Report Comparison). DISCERN is a significance-aware framework that weighs report errors based on their potential impact on patient care. Our results demonstrate that DISCERN powered by closed source LLMs aligns more closely with expert radiologist assessments than traditional metrics or current LLM evaluators, providing a more interpretable and clinically relevant benchmark. By modeling radiologist prioritization and entity-level feedback, DISCERN facilitates targeted model refinement and ensures the safer integration of generative AI into clinical workflows.

## Introduction

The demand for radiologic interpretation continues to rise, while radiologist workforce growth has not kept pace with the increasing volume and complexity of imaging examinations per patient^1,2^. Recent analyses of the U.S. radiologist workforce highlight persistent shortages, higher burnout rates, and increasing reporting delays, raising concerns about downstream impacts on patient care^3,4^. As imaging utilization expands across inpatient, outpatient, and emergency settings, scalable solutions that support radiologists while maintaining reporting quality are urgently needed.

Artificial intelligence (AI), particularly vision–language models (VLMs), has emerged as a promising approach to improving radiology workflow efficiency. Recent work demonstrates that VLMs capable of interpreting radiographs and generating draft reports can reduce documentation time up to 15% without compromising accuracy or readability^5^. Continued development and future clinical deployment of these models depend not only on model architecture and training data, but also on the availability of robust strategies to evaluate model performance at scale^6^. The clinical utility of these systems is directly linked to the use of clinically meaningful evaluation framework^7^. In practice, comprehensive manual evaluation of generated reports by radiologists is not feasible for large datasets, given the limited availability of expert time. As a result, scalable automated evaluation approaches are necessary to systematically assess report quality and guide model improvement.

Traditional automated natural language processing (NLP) metric, such as BLEU^8^, ROUGE^9^, METEOR^10^, BERTScore^11^, and RadGraph-F1^12^, primarily capture semantic alignment, lexical overlap, or entity-level similarity, but do not reflect clinical correctness. Indeed, high similarity scores can coexist with major diagnostic or localization errors^13^. To address these limitations, recent work has introduced structured, large-language-model (LLM)–based evaluation frameworks such as GREEN^13^, GEMAScore^14^, and SPEC-CXR^15^. These systems identify finding- or entity-level errors and categorize them into interpretable types, enabling more nuanced assessment than simple similarity metrics.

Despite these advances, existing framework treat discrepancies uniformly, without accounting for clinical significance. In contrast, radiologists prioritize findings by urgency, actionability, and impact on patient management. This mismatch limits their ability to reflect true clinical impact and may obscure clinically meaningful differences between models. Accordingly, there is a need for evaluation strategies that align with radiologists’ assessment and prioritization in routine practice.

In this work, we introduce DISCERN [Figure 1], a clinically grounded discrepancy aware evaluation framework for radiology report generation. DISCERN extends beyond existing approaches by (1) decomposing discrepancies into multiple structured categories, and (2) assigning a clinically meaningful significance score to each discrepancy. Our contributions are as follows:

**Figure 1.**
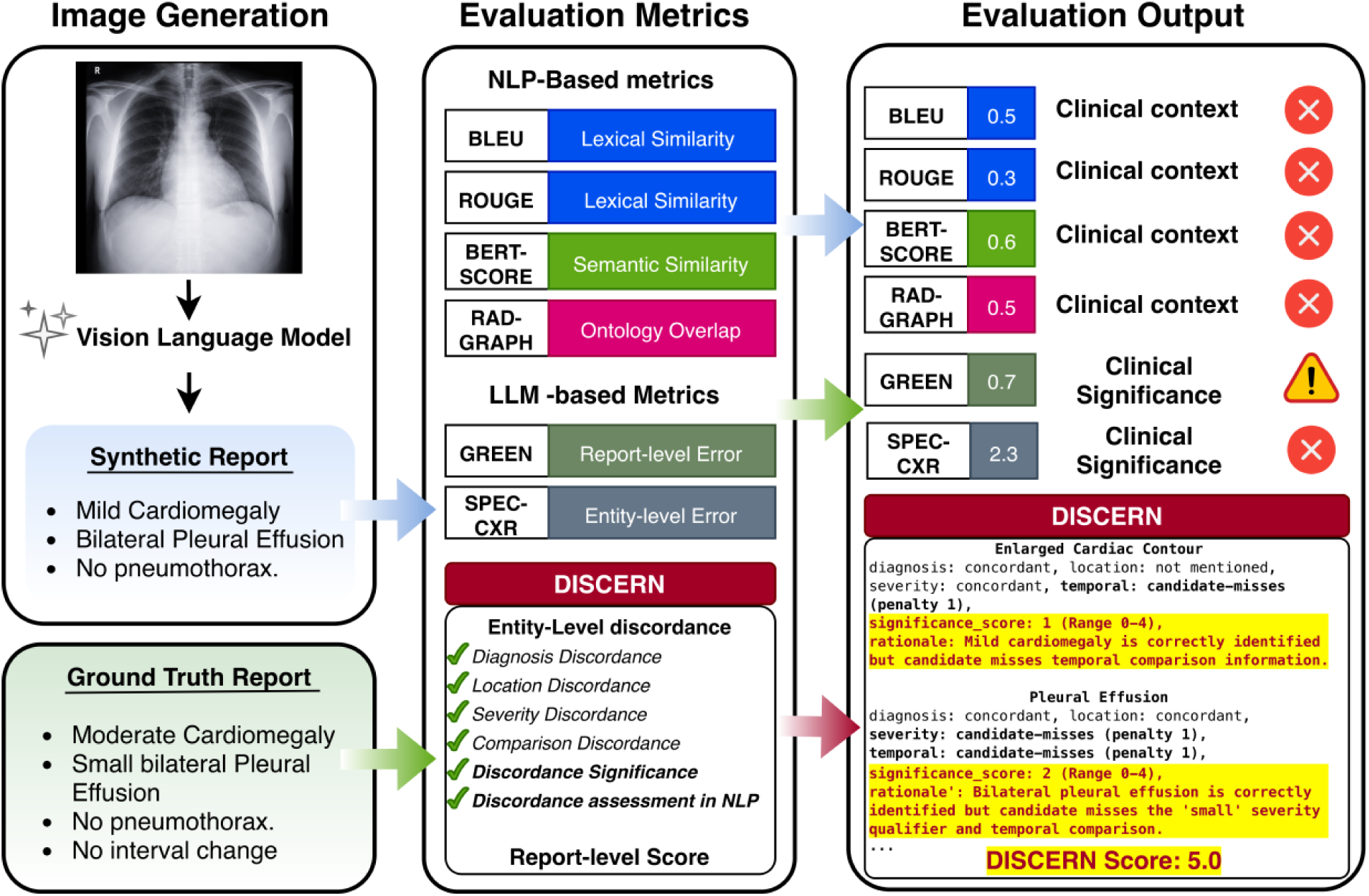
Comparison of NLP-based and LLM-based evaluation metrics with the proposed DISCERN framework. NLP metrics lack clinical contextual understanding, and existing LLM-based methods (e.g., SPEC-CXR^15^) do not quantify clinical significance. GREEN^13^ offers only coarse, two-level significance. DISCERN provides a five-level clinical-significance scale, interpretable explanations, and an integrated report-level score combining significance weighting and discordance penalties.

- A multi-attribute discrepancy taxonomy that separately evaluates diagnosis discordance and uncertainty; location, severity, and temporal discordance; and candidate additions or omissions.
- A clinically grounded significance scoring system (0–4) that ranks discrepancies by their real-world patient management impact and generates natural-language explanations.
- Extensive benchmarking of open-source and closed-source LLMs on two public radiology evaluation datasets ReXVal^16^ and RadEvalX^17^.

## Methods

### Overview of DISCERN

Our framework operates in four stages [Figure 2]. Given a reference (ground-truth) report *R* and a candidate (model-generated) report 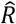, **Stage 1** extracts clinically meaningful entities from both reports using a pre-curated list of 121 radiologic entities^15^; **Stage 2** categorizes discrepancies for shared and missing entities across multiple attributes; **Stage 3**, assigns each entity-level discrepancy a 0–4 clinical significance score and generates a concise natural-language rationale explaining the assigned severity, and **Stage 4** assigns a penalty for each attribute discordance and calculates a significance-weighted sum of attribute penalties using the equation the following formulation:

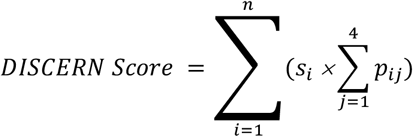

where *s*_*i*_ is the significance score of entity *i*; *p*_*ij*_ is the discordance penalty for attribute *j* of entity *i*; *n*= count of total identified entities, i.e. *count*(*entities*_*candidate*_ ∪ *entities*_*reference*_); and the count of attributes is 4 (diagnosis, location, severity, temporal).

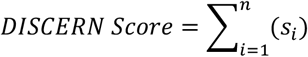

**Figure 2.**
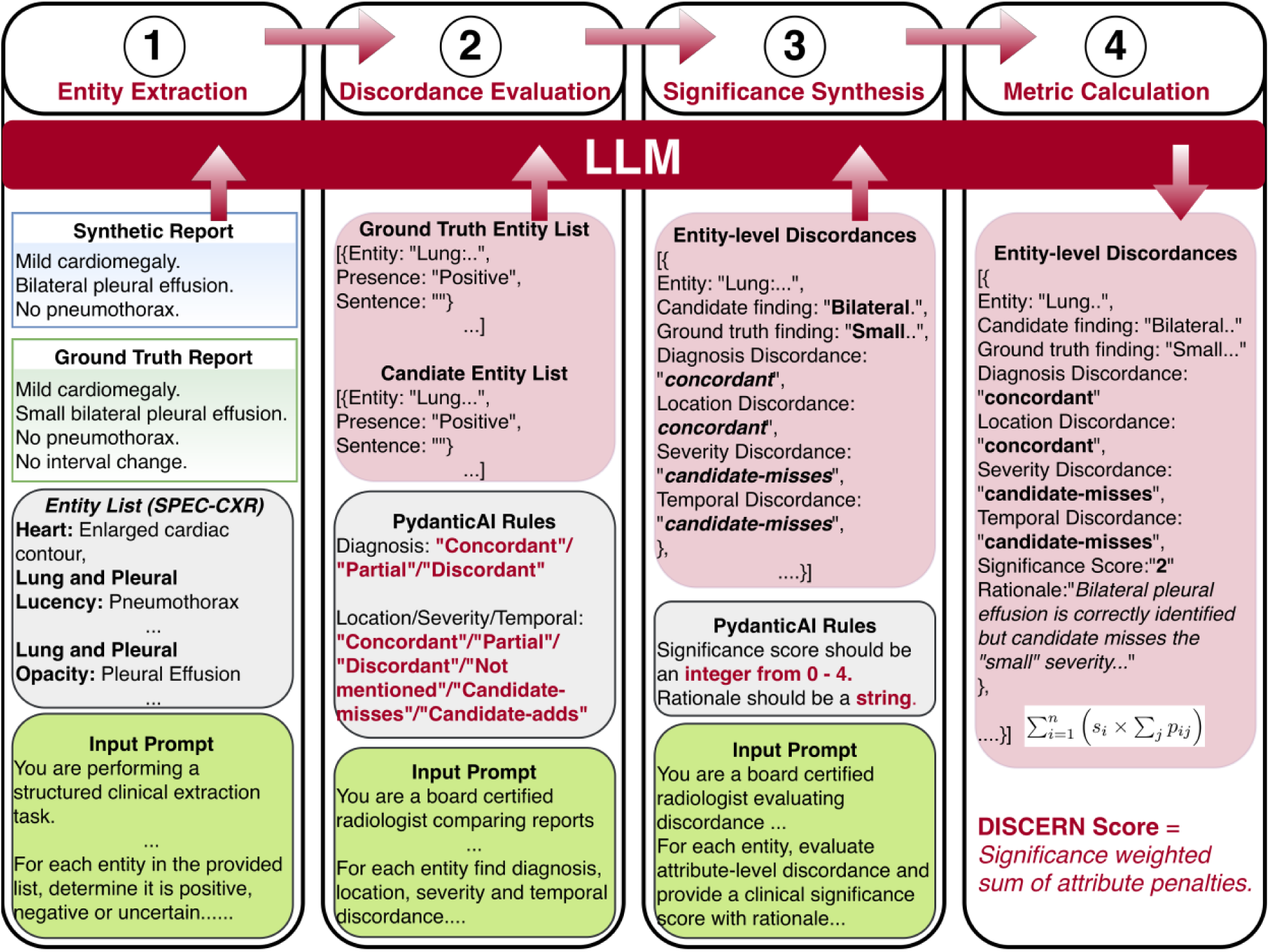
Overview of the DISCERN framework. In stage 1, a detailed input prompt is used with a pre-curated entity list^15^ to extract entities from both reports. In stage 2, entities present in both reports are assessed for attribute (*diagnosis, location, severity, temporal*)-level discordance. In stage 3, entity-level discordances are synthesized to generate clinical significance score along with a rationale. In stage 4, a weighted sum is calculated to generate DISCERN Score. (Red box (top box in stage 2, 3 and 4) is the output from their respective previous stage.)

### Entity List and Discrepancy Taxonomy

Entity List: Lee et al.^15^ curated and evaluated a comprehensive list of 121 entities to categorize all possible chest X-ray findings and diagnoses. Across 2,347 MIMIC-CXR reports, their entity set achieved 81.9% coverage, outperforming UMLS^18^ (73.7%) and CheXpert^19^ (56.8%). We used their entity list to evaluate proposed framework in this study.

### Discrepancy Taxonomy

We built a comprehensive taxonomy to identify discordance in the reports. For entities present in both reports, **presence** is categorized into three levels: *positive, negative*, or *uncertain*. For each entity specific evaluation, we decomposed discordance into four attributes: *diagnosis, location, severity*, and *temporal*. **Diagnosis** discordance is assessed at three levels for each shared entity: *concordant, partial*, or *discordant*. Whereas **location, severity**, and **temporal** change discordance is evaluated at 6 levels: *concordant, partial, discordant, not mentioned, candidate-adds*, or *candidate-misses*. **Clinical significance** per entity is scored on a 5-point rubric aligned with radiologist triage and management:

- **4**: time-sensitive discrepancy likely to change immediate management (*Critical*)
- **3**: likely to affect treatment or near-term diagnostic workup (*Important*)
- **2**: may influence follow-up or differential diagnosis (*Moderate*)
- **1**: incidental or chronic/stable discrepancy (*Low*)
- **0**: no meaningful impact or fully concordant entity (*None*).

We used *Pydantic* to enforce strict output schema validation. All generations satisfied the schema within a single retry.

## Experiments

### Datasets

*ReXVal*^16^ is a publicly available benchmark comprising expert radiologist evaluations of errors in synthetic chest X-ray reports. The dataset contains 200 pairs of reports, each assessed independently by six board-certified radiologists. For each pair, radiologists annotated six predefined error categories: false prediction, omission, incorrect location, incorrect severity, comparison addition, and comparison omission. For every error type, each rater recorded the number of clinically significant and clinically insignificant errors.

*RadEvalX*^17^ is a similarly constructed public evaluation dataset designed to assess radiology report generation models. It includes 100 report pairs evaluated by two board-certified radiologists. As in ReXVal, raters quantified the number of clinically significant and clinically insignificant errors per pair. In addition to the six error categories defined in ReXVal, RadEvalX expands the taxonomy to eight categories by incorporating uncertainty addition and uncertainty omission.

### Baseline comparison

We compared DISCERN against established NLP metrics, including BLEU, ROUGE, BERTScore, RadGraph-F1, and METEOR, as well as LLM-based evaluation frameworks such as GREEN and GEMAScore.

To assess the effect of backbone choice within DISCERN, we evaluated multiple large language models spanning a range of parameter scales and reasoning paradigms. These included both open-source and closed-source models, with closed-source systems deployed on secure institutional servers. The evaluated models were LLaMA-3.1-8B-Instruct, Gemma-3-12B, MedGemma-27B-it, DeepSeek-R1-Distill-QWEN-32B, GPT-OSS-120B, and the Claude family (Haiku 4.5, Opus 4.5, and Sonnet 4.6).

Performance was quantified by measuring correlation with radiologist annotations using Kendall’s τ and Spearman’s ρ coefficients. For ReXVal, correlations were computed with respect to the average rater annotations across the six board-certified radiologists. For each report pair, we used the mean count of clinically significant and clinically insignificant errors as the reference standard.

In contrast, RadEvalX provides consensus-level annotations following adjudication between two board-certified radiologists. Accordingly, correlations on RadEvalX were computed directly against these final agreement labels.

## Results

### Comparison with Baseline Metrics

Figure 3 shows Kendall’s τ and Spearman’s ρ correlations with radiologist annotations on ReXVal and RadEvalX. Points above the diagonal indicate stronger alignment with clinically significant than insignificant errors—reflecting appropriate prioritization—while strong performance on both axes indicates overall agreement with radiologist judgments.

**Figure 3.**
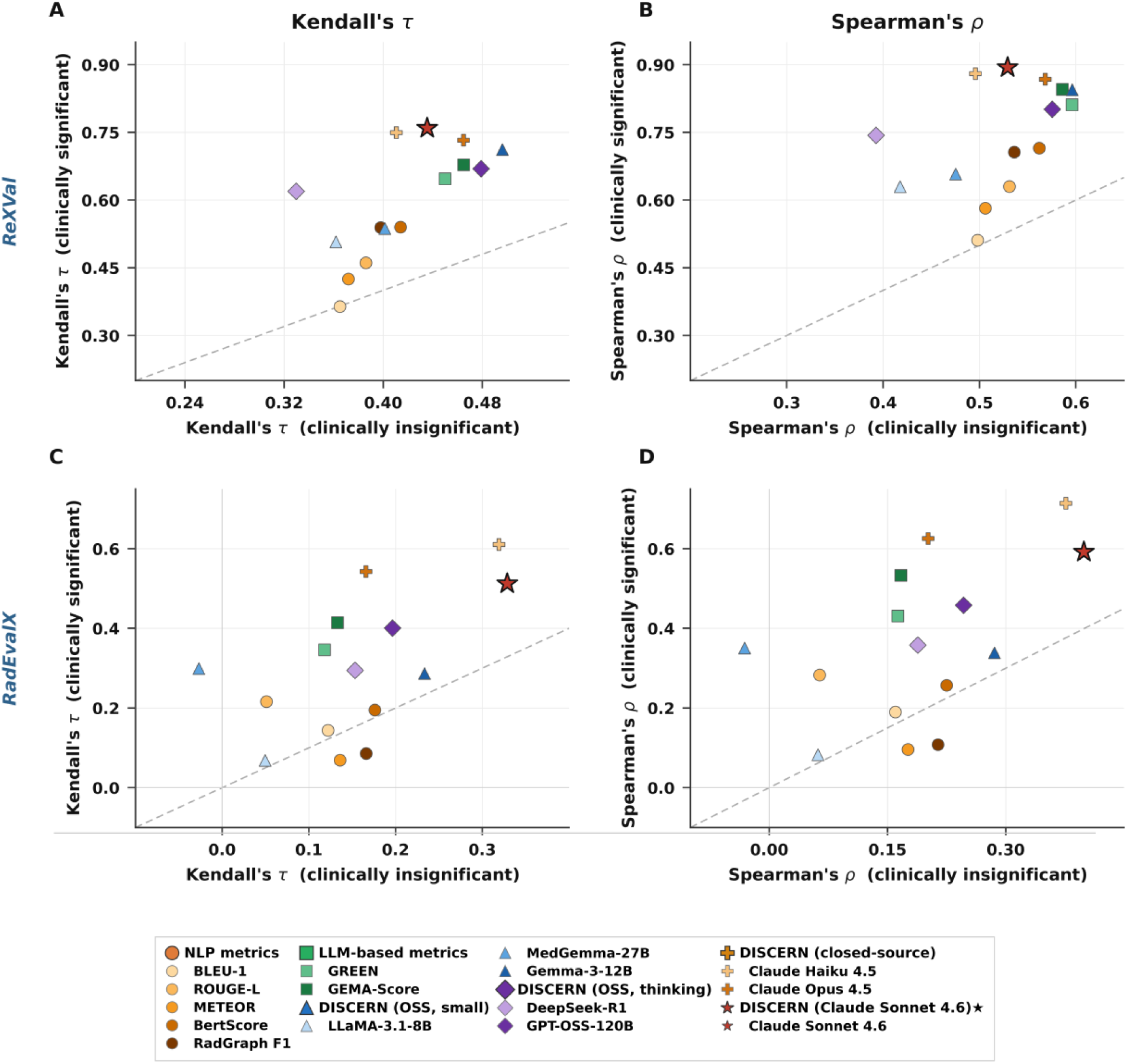
Correlation of automated metrics with radiologist annotations across ReXVal and RadEvalX datasets. Kendall’s τ (left) coefficient and Spearman’s ρ coefficient (right) are shown for clinically insignificant (x-axis) and clinically significant (y-axis) errors.

#### ReXVal

NLP-based metrics demonstrated modest performance and clustered near the diagonal (*τ*_*sig*_: 0.364 - 0.540), suggesting limited sensitivity to clinical importance. RadGraph-F1 and BERTScore performed best among this group but showed only moderate correlations overall. LLM-based baselines substantially improved alignment. GREEN (*τ*_*sig*_= 0.647, *ρ*_*sig*_= 0.811) and GEMA-Score (*τ*_*sig*_= 0.678, *ρ*_*sig*_= 0.845) achieved strong correlations on clinically significant errors while also demonstrating robust agreement on insignificant discrepancies (*τ*_*sig*_= 0.450 and 0.465, respectively). Although both axes increased in parallel, they exhibited slight preferential weighting toward clinically meaningful errors.

DISCERN (with Claude-Sonnet-4.6 backbone) achieved the highest overall correlations *τ*_*sig*_= 0.725, *ρ*_*sig*_= 0.858), improving over GEMA-Score by +0.047 in *τ*_*sig*_ and +0.013 in *ρ*_*sig*_. Importantly, it maintained strong alignment on insignificant errors *τ*_*insig*_= 0.506, *ρ*_*insig*_= 0.623), indicating improved overall agreement while preserving prioritization of clinically significant discrepancies.

#### RadEvalX

RadEvalX represents a less commonly used and distributionally distinct dataset, and all metrics exhibited reduced performance relative to ReXVal. NLP-based approaches showed attenuated correlations *τ*_*sig*_: 0.069 – 0.216; *ρ*_*sig*_: 0.096–0.283), indicating limited robustness under cross-dataset evaluation. LLM-based baselines also experienced notable declines. GEMA-Score decreased from *τ*_*sig*_ = 0.678 → 0.414 (*ρ*_*sig*_: 0.845 → 0.533), and GREEN dropped from *τ*_*sig*_= 0.647 → 0.346 (*ρ*_*sig*_: 0.811 → 0.431). These reductions may reflect distributional differences between datasets, including variations in reporting style, clinical content, and evaluation protocol. In contrast, DISCERN (Claude-haiku-4.5) demonstrated stronger cross-dataset generalization, achieving *τ*_*sig*_= 0.611 and *ρ*_*sig*_= 0.714.

Despite the overall performance attenuation observed across methods, DISCERN maintained substantially higher alignment with radiologist judgments than both NLP and LLM baselines, suggesting improved robustness to dataset variation.

### Effect of Backbone Model Choice

Performance stratified by model family and scale. Smaller open-source models (LLaMA-3.1-8B, MedGemma-27B) underperformed across datasets (ReXVal *τ*_*sig*_=0.507 and 0.537), well below top closed-source models (*τ*_*sig*_ ≥ 0.732). Mid-size open models were competitive on ReXVal (Gemma-3-12B: *tau*_*sig*_=0.712; Qwen3-30B:*τ*_*sig*_= 0.655) but degraded substantially on RadEvalX (0.287 and 0.200), indicating sensitivity to distributional shift. DeepSeek-R1 showed reasonable significant-error alignment on ReXVal (*τ*_*sig*_=0.619) with lower insignificant-error correlation (*τ*_*insig*_=0.330), but similarly declined on RadEvalX (*τ*_*sig*_=0.295).

Closed-source Claude models were most consistent. On ReXVal, significant-error alignment increased with scale (Haiku-4.5: 0.749 → Sonnet-4.6: 0.759), with strong insignificant-error correlations (0.411–0.465). On RadEvalX, performance decreased overall but remained highest among Claude variants (Haiku-4.5: *τ*_*sig*_=0.611), with minor reordering across scales.

### Dual Sensitivity and Generalization

A clinically meaningful metric must capture both significant errors (for ex. actionable findings, acute symptoms, patient safety etc.) and insignificant errors (for ex. non-critical misses, chronic symptoms, report completeness etc.). Across datasets, top DISCERN configurations improved significant-error alignment without sacrificing insignificant-error correlation, indicating additive gains rather than trade-offs.

Baseline LLM methods showed larger cross-dataset drops (GREEN: 0.647 → 0.346; GEMA-Score: 0.678 → 0.414), consistent with distributional variation. In contrast, DISCERN maintained strong significant-error alignment on RadEvalX (*τ*_*sig*_=0.611) with competitive insignificant-error correlation (*τ*_*insig*_=0.319), demonstrating improved robustness.

Insignificant-error alignment remained modest across methods on RadEvalX, likely reflecting heterogeneous incidental findings and limited fine-grained supervision—highlighting the need for severity-aware training data in future work.

## Conclusions and future work

We presented DISCERN, a clinical significance–aware framework for radiology report evaluation that decomposes discrepancies into structured attributes and assigns entity-level severity scores (0–4). By explicitly prioritizing clinically actionable findings rather than treating all discrepancies uniformly, DISCERN provides a more clinically aligned evaluation signal.

Across two independent datasets and multiple backbone LLMs, DISCERN consistently improved alignment with radiologist judgments on clinically significant errors while maintaining competitive performance on insignificant discrepancies. Strong closed-source models achieved the highest overall performance, though mid-size open-source models showed encouraging results, suggesting that scalable deployment within this framework is feasible.

However, performance on insignificant-error assessment remained modest across all models, including the strongest closed-source systems. This consistent gap may reflect the inherent difficulty of assessing low-salience discrepancies. It may indicate current limitations in how LLMs internalize nuanced clinical prioritization beyond surface-level semantic concordance.

Several additional limitations warrant consideration. Clinical significance ideally depends on indication, history, and contextual parameters that were not available here. Dataset-specific differences in error taxonomies and annotation strategies may also influence results. While DISCERN employs a comprehensive attribute schema, practical constraints required adapting the taxonomy to each dataset, which may have introduced simplifications. Finally, reliance on large closed-source models entails non-trivial computational and financial costs.

Future work should focus on developing richer, context-aware datasets with fine-grained severity supervision, as well as smaller, robust models capable of granular clinical reasoning. Advancing nuanced insignificant-error assessment will be essential for truly comprehensive report evaluation. As generative radiology systems move closer to clinical deployment, clinically grounded, generalizable, and efficient evaluation frameworks such as DISCERN will be critical to ensuring that measured improvements translate into meaningful gains in patient care.

## Data Availability

All data used for the evaluation is available online at https://physionet.org/content/rexval-dataset/1.0.0/ and https://physionet.org/content/rad-eval-x/1.0.0/. Codebase is available at https://github.com/rakshrma/discern/.

